# Global Infodemiology of COVID-19: Focus on Google Web Searches and Instagram Hashtags

**DOI:** 10.1101/2020.05.21.20108910

**Authors:** Alessandro Rovetta, Akshaya Srikanth Bhagavathula

**Author notes:** **Corresponding author**: Alessandro Rovetta, Mensana srls, Research and Disclosure Division, Via Moro Aldo 5 – 25124 Brescia, Italy.

## Abstract

**Background:** Though ‘infodemiological’ methods have been used in COVID-19 research, an examination of the extent of infodemic monikers (misinformation) use on the Internet remains limited.

**Objective:** To investigate Internet search behavior related to COVID-19 and examine the circulation of infodemic monikers through two platforms—Google and Instagram—during the current global pandemic.

**Methods:** Using Google Trends and Instagram hashtags (#), we explored Internet search activities and behaviors related to the COVID-19 pandemic from February 20, 2020, to May 06, 2020. We investigated the names used to identify the virus, health and risk perception, life during the lockdown, and information related to the adoption of COVID-19 infodemic monikers. We computed the average peak volume (APC) with a 95% confidence interval (CI) during the study period for the monikers.

**Results:** The top five COVID-19-related terms in the Google searches were “coronavirus”, “corona”, “COVID”, “virus”, “corona virus”, and “COVID-19”. Countries with a higher number of COVID-19 cases had a higher number of COVID-19 queries on Google. The monikers “coronavirus ozone”, “coronavirus laboratory”, “coronavirus 5G”, “coronavirus conspiracy” and “coronavirus bill gates” were widely circulated on the Internet. Searches about ‘tips and cures’ for COVID-19 spiked in relation to the U.S. president speculating about a ‘miracle cure’ and suggesting the injection of disinfectant to treat the virus. Around two-thirds (66.1%) of Instagram users used the hashtags “COVID-19”, and “coronavirus” to disperse virus-related information.

**Conclusion:** Globally, there is a growing interest in COVID-19, and numerous infodemic monikers continue to circulate on the Internet. Based on our findings, we hope to encourage mass media regulators and health organizers to be vigilant and diminish the use and circulation of these infodemic monikers on the Internet, to decrease the spread of misinformation.

## Introduction

Globally, the Internet is an extremely important platform for obtaining knowledge and information about the novel coronavirus (COVID-19) pandemic [1-3]. The Google Trends tool provides real-time insights into internet search behavior on various topics, including COVID-19 [4]. Social media platforms such as Facebook, Twitter, and Instagram allow users to communicate their thoughts, feelings, and opinions by sharing short messages. A unique aspect of social media data from Instagram is that image-based posts are accessible, and the use of topic-related hashtags (#) allows access to topic-related information for all Internet users [5]. In general, there is a growing interest in examining social data to understand and monitor public behavior in real-time [6,7].

Research on the Internet and social data are called *Infodemiology* or *Infoveillance* studies [8]. *Infodemiology* is defined as “the science of distribution and determinants of information in an electronic medium, specifically the Internet, or in a population, with the ultimate aim to inform public health and public policy” [9]. Although several studies have been conducted using ‘infodemiological’ methods in COVID-19 research, a limited number of studies have examined the extent of COVID-19-related misinformation on the Internet [10-14]. The fake news, misleading, and misinformation circulating on the Internet are referred to as “infodemic monikers”. These monikers can profoundly affect public health communication and also contribute to xenophobia [12-17]. “*Infodemic monikers*” are defined as substantially erroneous information, which give rise to interpretation mistakes, fake news, episodes of racism, or any other forms of misleading information circulating on the internet [14]. In this context, we aimed to investigate the Internet search behavior related to COVID-19 and the extent of infodemic monikers circulating on Google and Instagram during the current pandemic period in the world.

## Methods

We used Google Trends and Instagram hashtags to explore internet search activities and behaviors related to the COVID-19 pandemic from February 20, 2020, to May 06, 2020. We investigated the following: names used to identify the virus, health and risk perception, life during the lockdown, and information related to the adoption of infodemic monikers related to COVID-19. The complete list terms used to identify the most frequently searched queries in Google and the hashtag suggestions for Instagram are presented in Supplementary File 1.

The obtained infodemic monikers are characterized as follows:

1. *Generic:* The moniker confuses, due to lack of specificity.
2. *Misinformative*: The moniker associates a certain phenomenon with fake news.
3. *Discriminatory:* The moniker encourages the association of a problem with a specific ethnicity and/or geographical region.
4. *Deviant:* The moniker does not identify the requested phenomenon.
5. *Other specificities:* We keep two additional points for special cases that prove exceptionally serious.

To determine the severity of various infodemic monikers circulating on the Internet, each moniker was assigned 1 to 2 points on the infodemic scale (I-scale) ranging from 0 (minimum) to 10 (maximum). Based on the sum of the I-scale scores, the infodemic monikers were classified as follows:

- Not infodemic: 0
- Lowly infodemic: 1
- Moderately infodemic: >1-4
- Highly infodemic: 5-8
- Extremely infodemic: 9-10

For each search keyword considered, Google Trends provided normalized data in the form of relative search volume (RSV) based on search popularity ranging from 0 (low) to 100 (highly popular). Using these RSV values, we computed the average peak volume (APC) with a 95% confidence interval (CI) during the study period.

Instagram, a platform for image-based posts with hashtags (#) was screened. We retrieved content based on hashtags through image classifiers, every 3-4 days during the study period. All irrelevant content was excluded. The data collected included contents posted on Instagram and self-reported user demographic information. No personal information, such as emails, phone numbers, or addresses, was collected. The data from the Instagram hashtags were collected manually, through the Instagram-suggested tags associated with specific countries.

All data used in the study were obtained from anonymous open sources. Thus, ethical approval was not required.

## Results

The top five COVID-19 related infodemic and scientific terms used in Google searches were “coronavirus”, “corona”, “COVID”, “virus”, “corona virus”, and “COVID-19” [Figure 1]. The most frequently used keywords globally were “coronavirus” (APC: 1378, 95% CI: 1246-1537), followed by “corona” (APC: 530, 95% CI: 477-610) and “COVID” (APC: 345, 95% CI: 292-398). Several keywords related to COVID-19 (Table 1) were identified, of the top 10, five had an I-scale value of 8: “corona”, “corona Italy”, “corona Deutschland”, “corona China” and “corona Wuhan”.

**Table 1:** Top infodemic and scientific Google searches related to COVID-19 in the world

| **Keyword** | **APC** | **95% CI** | **I-scale value** |
| --- | --- | --- | --- |
| coronavirus | 1378 | 1246 – 1537 | 3 |
| corona | 530 | 477 – 610 | 8 |
| COVID | 345 | 292 – 398 | 1 |
| virus | 239 | 212 – 292 | 7 |
| corona virus | 159 | 133 – 186 | 6 |
| coronavirus Italy | 54 | 45 – 62 | 4 |
| COVID-19 | 53 | 45 – 60 | 0 |
| coronavirus USA | 32 | 29 – 36 | 4 |
| coronavirus China | 30 | 25 – 34 | 6 |
| coronavirus Germany | 23 | 20 – 27 | 4 |
| corona Italy | 13 | 12 – 14 | 8 |
| corona Deutschland | 12 | 10 – 14 | 8 |
| SARS | 9 | 8 – 10 | 6 |
| corona China | 9 | 7 – 11 | 8 |
| corona Wuhan | 1 | 0 – 2 | 8 |
| SARS-CoV-2 | 1 | 0 – 1 | 0 |

Queries in APC: average peak volume; CI: confidence interval; I-scale: infodemic scale ranging from 0-10

**Figure 1:**
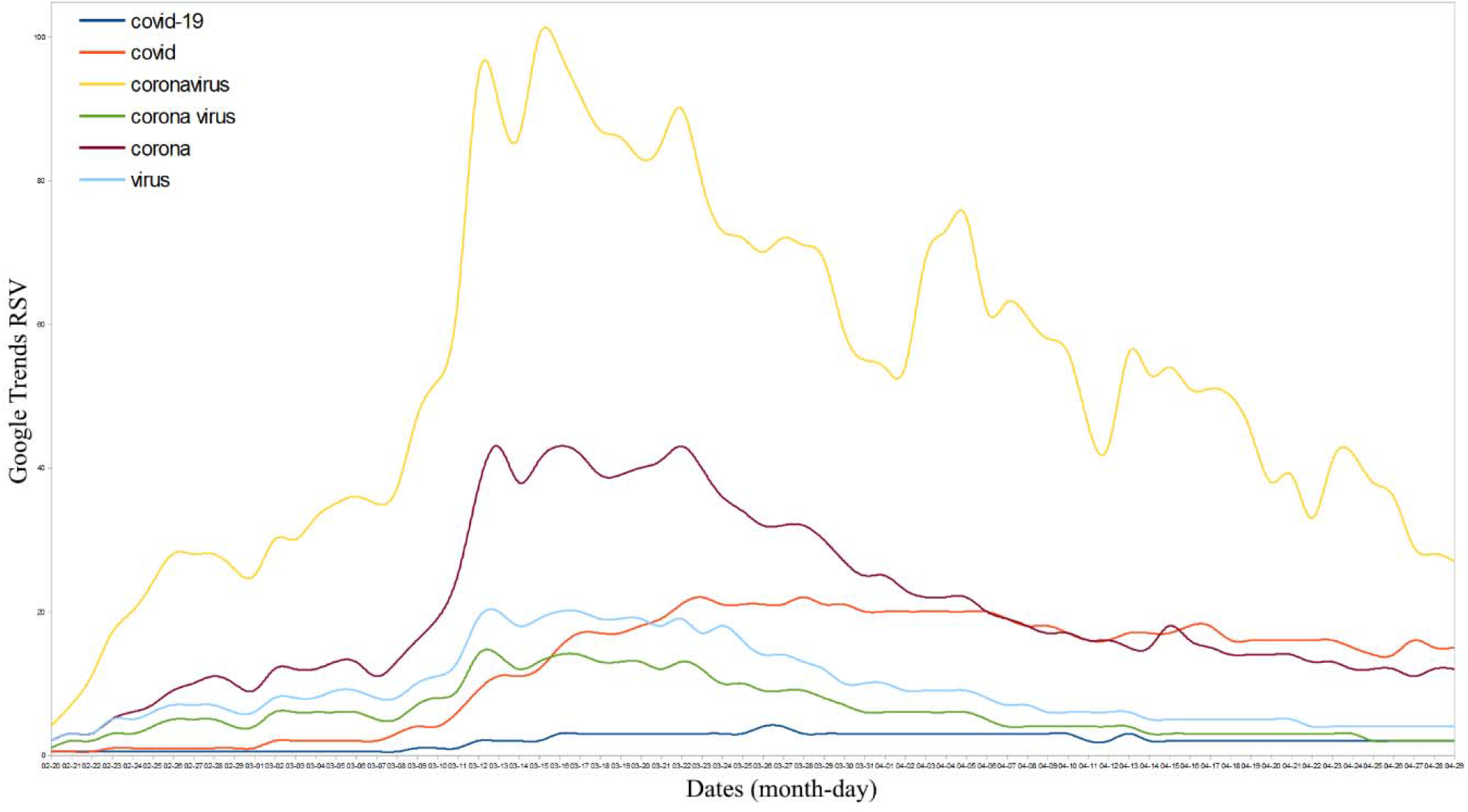
Top global scientific and infodemic names related to COVID-19 in the Google

The country-wise dispersion of the scientific and infodemic names of COVID-19 used in Google searches are shown in Figure 2. Countries with a higher number of COVID-19 cases per 1 million population have recorded greater Google search queries related to COVID-19 (Italy, Spain, Ireland, Canada, France, and Qatar). These COVID-19-related search queries were significantly correlated with the incidence of COVID-19 cases across the countries (Pearson R = 0.45, *p*<0.05).

**Figure 2:**
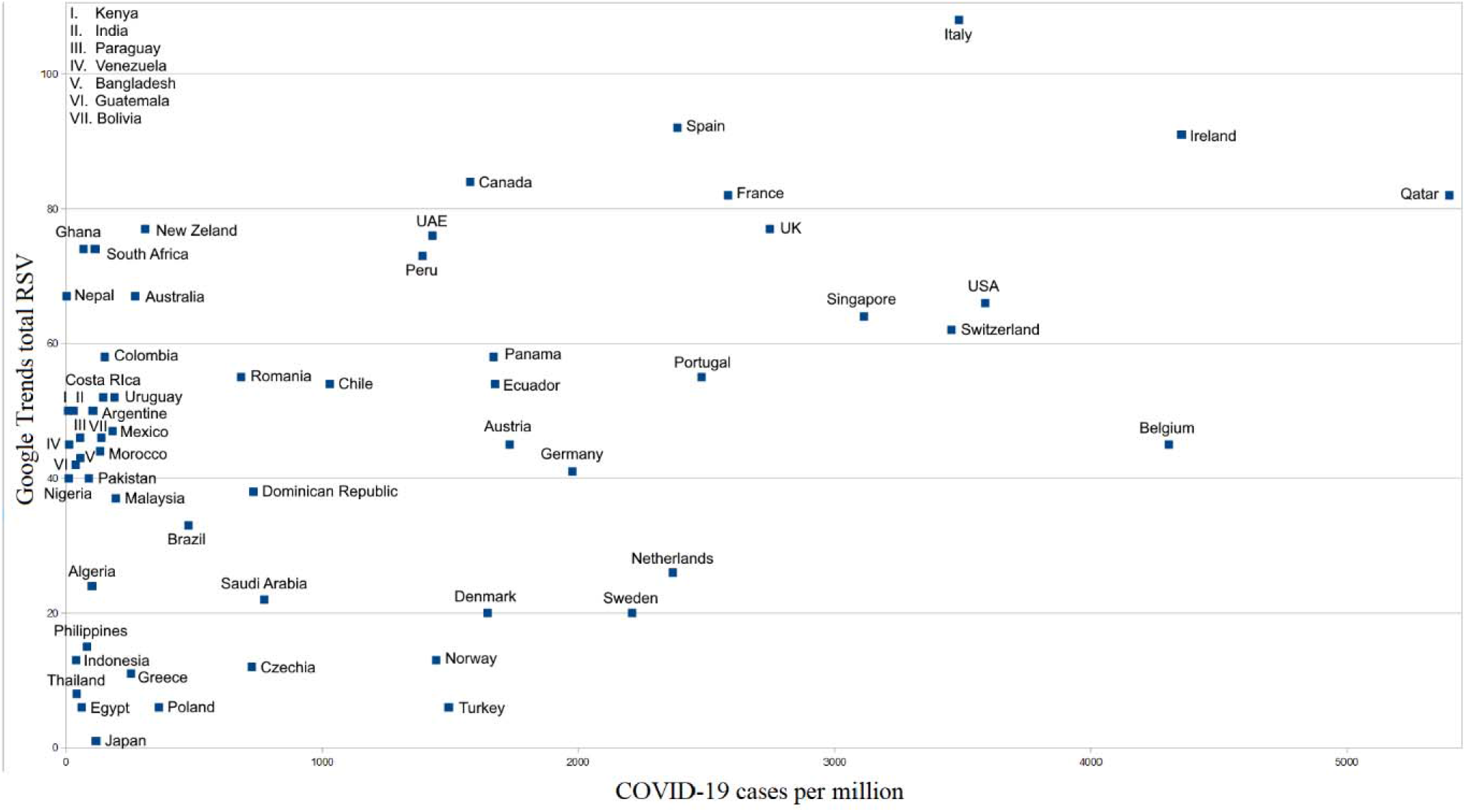
Countries-wise dispersion of scientific and infodemic names of COVID-19

The top COVID-19-related infodemic monikers such as “coronavirus ozone”, “coronavirus laboratory”, and “coronavirus 5G” frequently circulated on the Internet are presented in Table 2. The following are infodemic monikers with the highest I-scores globally, “coronavirus conspiracy” (I-score: 10), “coronavirus laboratory” (I-score: 9), and “coronavirus 5G” (I-score: 9). Additionally, the use of monikers with moderate to high infodemicity far exceeded the use of scientific names (Table 2): 57% of Google web searches are moderately infodemic (total APC: 109, 95% CI: 89 – 139) and 16% highly infodemic (total APC: 30, 95% CI: 25 – 34). The circulation of these infodemic monikers was further examined to understand the events associated with these searches. Infodemic monikers related to coronavirus origins, such as SARS-CoV-2 made in the laboratory” went viral (APC: 41) when the National Association Press Agency (NAPA) from Italy posted a 2015 video about the origins of SARS-CoV-2 virus on March 25, 2020[18]. In addition, the moniker reached the breakout level (RSV: 100) on April 17, 2020, when the French Noble Prize winner Prof. Luc Montagnier stated that the new coronavirus is the result of a laboratory accident in the Wuhan high-security laboratory in China [19]. Detailed information on the different infodemic monikers and the associated events are shown in Figure 3.

**Table 2:** Top global infodemic Google searches related to COVID-19

| **Keyword** | **APC** | **95% CI** | **I-scale value** |
| --- | --- | --- | --- |
| coronavirus ozone | 19 | 15 – 22 | 6 |
| coronavirus laboratory | 16 | 12 – 19 | 9 |
| coronavirus 5G | 10 | 8 – 13 | 9 |
| coronavirus conspiracy | 9 | 8 – 11 | 10 |
| coronavirus bill gates | 8 | 7 – 10 | 6 |
| coronavirus milk | 7 | 6 – 8 | 6 |
| coronavirus military | 4 | 4 – 5 | 8 |
| coronavirus uv | 3 | 3 – 4 | 8 |

Queries in APC: average peak volume, and CI: confidence interval; I-scale: infodemic scale ranging from 0-10

**Figure 3:**
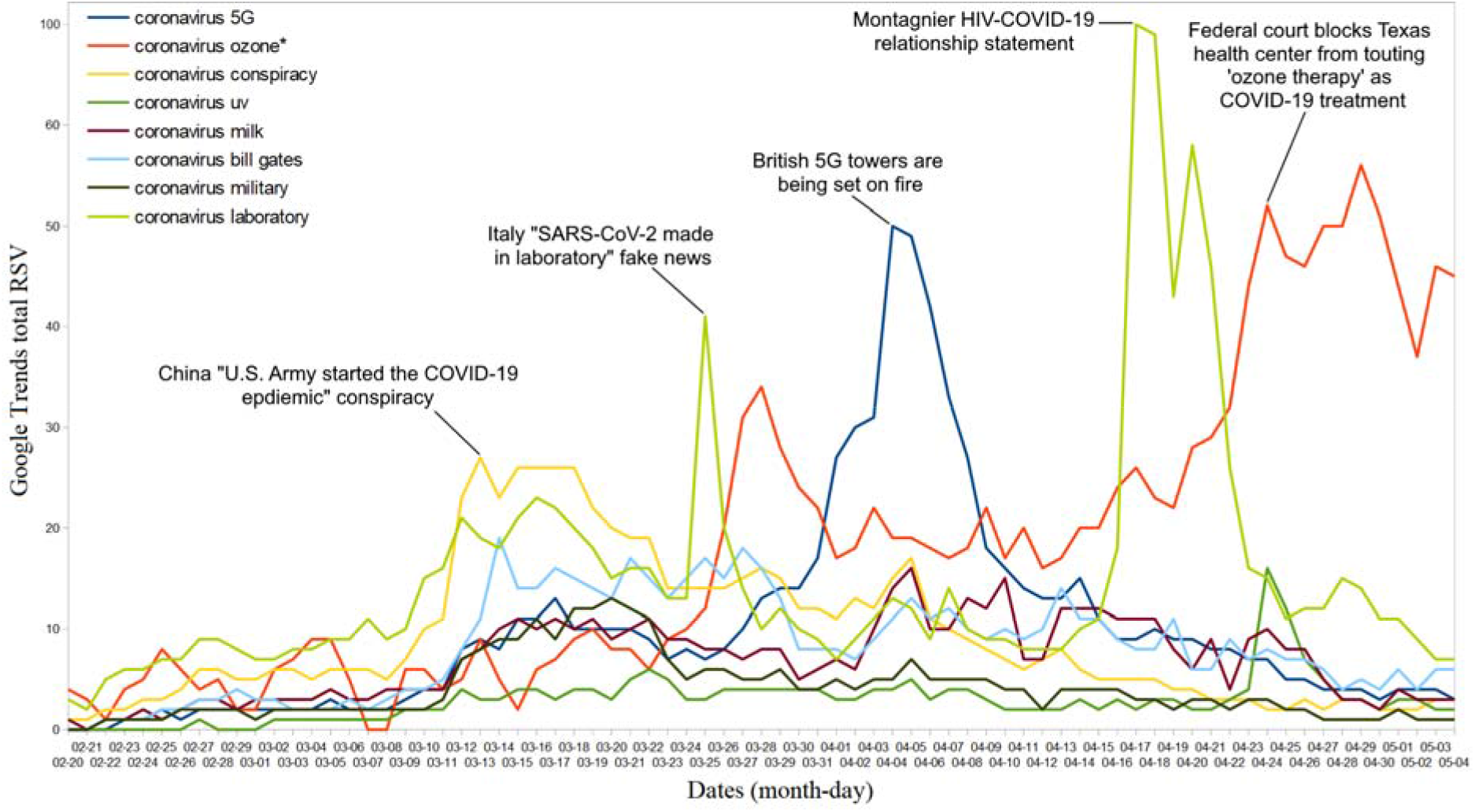
Top high and extreme infodemic global web searches related to COVID-19. * the ozone-coronavirus association concerns both the alleged therapy against COVID-19 and the stratospheric phenomenon. Although the second association is not directly infodemic, it can contribute to the spread of the first.

The top searches related to health, precautions, and COVID-19 news are presented in Figure 4. Google searches related to COVID-19 news remained at the top throughout the pandemic period. However, searches related to ‘tips and cures’ for COVID-19 spiked multiple times when the U. S. president suggested that hydroxychloroquine (an unproven drug) was a ‘miracle cure’ on April 4, 2020 (RSV: 70) [20] and also injecting disinfectant to treat COVID-19 on April 24, 2020 (RSV: 53) [21]. Other searches related to the use of medical masks and disinfectants (APC: 23, 95% CI: 21 – 25), lockdown (APC: 19, 95% CI: 16 – 22), and COVID-19 symptoms (APC: 12, 95% CI: 10 – 15), are less frequently used in Google searches.

**Figure 4:**
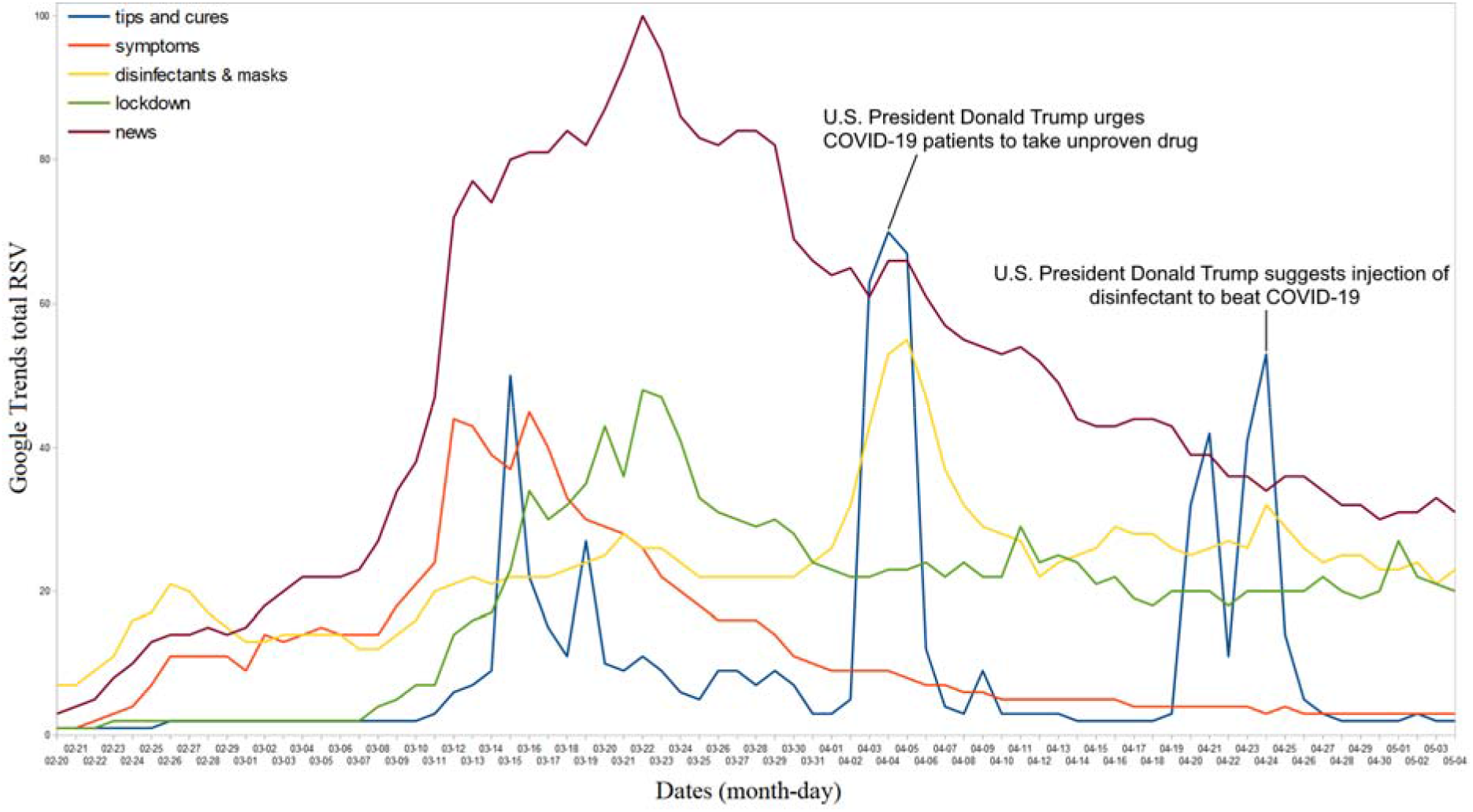
Top global web searches related to health, precautions and COVID-19 news.

The top 10 COVID-19-related hashtags used on Instagram (country-specific) and groups and topics associated with these hashtags are summarized in Table 3. Around one million users from Italy used ‘covid-19’ as a hashtag 3.6 million times to communicate information related to health, stay-home/safe (93.3 million times). These hashtags remained at the top for use for COVID-19-related communication on Instagram. Similarly, Instagram users from Brazil (551,000), Spain (376,000), Indonesia (298,000), and other countries were mostly used Instagram frequently to distribute COVID-19 related information. Moreover, the contribution of the ‘covid-19’ hashtag for COVID-19 related information was 35.6%, followed by ‘coronavirus’ (30.5%), ‘corona’ (25.6%), and ‘COVID’ (8%) [Figure 5].

**Table 3:** Top 10 Instagram hashtags related to COVID-19

| Top 10 COVID-19-related Instagram hashtags (#) |  |  |  |  |  |  |
| --- | --- | --- | --- | --- | --- | --- |
| *Rank* | *Country group* | *Quantity<sup>†</sup>* | *Hashtag group* | *Quantity<sup>†</sup>* | *Hashtag topic group* | *Quantity<sup>†</sup>* |
| 1 | Italy | 9.63 | covid-19 | 306 | Health-stay home/safe | 933 |
| 2 | Brazil | 5.51 | coronavirus | 267 | lockdown life | 718 |
| 3 | Spain | 3.76 | corona | 188 | masks | 135 |
| 4 | Indonesia | 2.98 | covid | 69 | memes | 25 |
| 5 | Turkey | 2.44 | corona memes | 14 | gym/fitness | 24 |
| 6 | India | 1.65 | coronavirus Italy | 9.63 | art/hobbies | 22 |
| 7 | Malaysia | 0.89 | coronado | 8.19 | cooking | 21 |
| 8 | Dominican Republic | 0.83 | corona time | 7.12 | fashion | 16 |
| 9 | USA | 0.75 | coronavirus memes | 6.41 | hair/beard style | 14 |
| 10 | Argentina | 0.74 | coronavirus Brazil | 5.51 | fun/party | 13 |

Searches identified until: May 6, 2020; <sup>†</sup> multiples in 100,000

**Figure 5:**
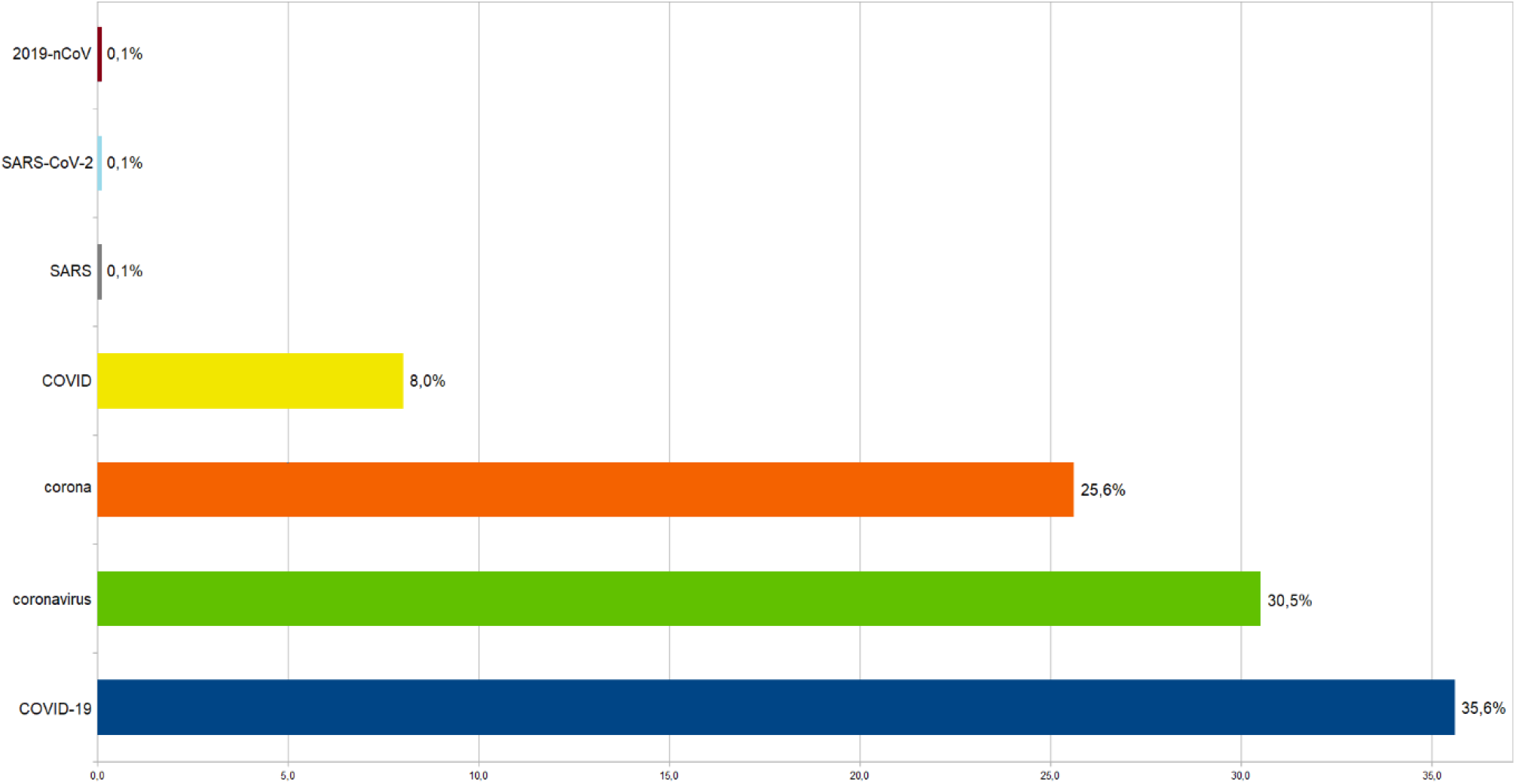
Top Instagram hashtags related to COVID-19 scientific and infodemic names.

## Discussion

In light of the ongoing COVID-19 pandemic, we are the first to investigate the Internet search behavior of the public and the extent of Infodemic monikers circulated on Google and Instagram globally. Our results suggest that (i). “coronavirus”, “corona”, “COVID”, “virus”, “corona virus”, and “COVID-19” are the top five terms used in the Google searches. (ii). countries (e.g., Italy, Spain, Ireland, Canada, and France) with a high incidence of COVID-19 cases (per million) have recorded greater Google search queries about COVID-19. (iii). “coronavirus ozone”, “coronavirus laboratory”, “coronavirus 5G”, “coronavirus conspiracy” and “coronavirus bill gates” are widely used infodemic monikers on the Internet. However, the “coronavirus conspiracy” was the only moniker that achieved the highest I-score of 10. (iv). though COVID-19 news remains at the top, web searches related to ‘tips and cures’ for COVID-19 spiked when the U.S. president speculated about a ‘miracle cure’ and the injection of a disinfectant to treat COVID-19. (v). Around two-thirds (66.1%) of Instagram users use “COVID-19”, and “coronavirus” as a hashtag to disperse the information related to COVID-19.

Exploring research using nontraditional data sources such as social media has several implications. First, our results demonstrated a potential application for using Instagram as a complementary tool to aid in understanding online search behavior and also provided real-time tracking of infodemic monikers circulated on the Internet. The strength of this study is the investigation of various infodemic monikers dispersed on the internet and correlating them with the events associated with that particular day. By characterizing and classifying various infodemic monikers based on the degree of infodemicity scores (I-score), researchers can foster new methods of using social media data to monitor the monikers’ outcomes. The analysis and methods used in this study could leverage public health and communication agencies in identifying and diminishing infodemic monikers circulating on the Internet.

Findings from this study validate and extend previously published works that used Google keywords [1,12,13]. We also demonstrate the potential for the use of Instagram hashtags to monitor and predict both the cyber behavior and relaying of misinformation on the Internet [22-24]. In 2017, *Guidry et al*. studied Ebola-related risk perception in Instagram users and identified that a significant proportion of posts on Instagram had rampant misinformation about the Ebola disease during the outbreak [22]. In addition, the percentage of Instagram posts and tweets posted by health organizations (CDC, WHO, MSF) to correct misinformation are less than 5% [22]. In general, negative information posted on the Internet tends to receive a greater weight among netizens. Thus, this should be counter-balanced with evidence-based solution content from health organizations, particularly in the current pandemic situation. For example, when the US president suggested injecting disinfectant to treat COVID-19, the number of Google searches considering it as a cure sharply increased (APC:53) and also implicated 30 cases of disinfectant poisoning within 18 hours in New York City [25]. Health authorities should be vigilant and provide more positive and informative messages to combat the circulation of infodemic monikers on social media. Future studies will need to investigate the influence of infodemic monikers on individual cyber behavior.

## Limitations

Our study used Google Trends, which provides the search behavior of people using their search engines, but not others. We mainly focused on Google and Instagram for data retrieval. Future studies should consider studying the same topic on other social media platforms to capture a more diverse population of users. Instagram searches were conducted manually, introducing a variable of error. Going forward, the use of an automated program can improve the accuracy of the data collected and analyzed. Lastly, Google Trends did not provide any information about the methods used to generate search data and algorithms.

## Conclusion

Using Google Trends and Instagram hashtags, the present study identified that there is a growing interest in COVID-19 globally and in countries with a higher incidence of the virus. Searches related to ‘COVID-19 news’ are quite frequent and two-thirds (66.1%) of Instagram users have used “COVID-19”, and “coronavirus” as hashtags to disperse information related to the virus. Several infodemic monikers are circulating on the Internet, with “coronavirus conspiracy” identified as the most popular moniker (I-score of 10). Given the prevalence of the infodemic moniker use, mass media regulators and health organizers should monitor and diminish the impact of these monikers. These governing bodies should also be encouraged to take serious actions against those spreading misinformation in social media.

## Data Availability

All external data used for research can be found in the references reported in the paper.

## Acknowledgment

None

## Data availability

All the data related to this study are presented in the Supplementary file.

